# GGC expansion in *ZFHX3* causes SCA4 and impairs autophagy

**DOI:** 10.1101/2023.10.26.23297560

**Authors:** Karla P. Figueroa, Caspar Gross, Elena Buena Atienza, Sharan Paul, Mandi Gandelman, Tobias Haack, Naseebullah Kakar, Marc Sturm, Nicolas Casadei, Jakob Admard, Joohyun Park, Christine Zühlke, Yorck Hellenbroich, Jelena Pozojevic, Saranya Balachandran, Kristian Händler, Simone Zittel, Dagmar Timmann, Friedrich Erdlenbruch, Laura Herrmann, Thomas Feindt, Martin Zenker, Claudia Dufke, Jeannette Hübener-Schmid, Daniel R. Scoles, Arnulf Koeppen, Stephan Ossowski, Malte Spielmann, Olaf Riess, Stefan M. Pulst

## Abstract

Despite linkage to 16q in 1996, the mutation for spinocerebellar ataxia type 4 (SCA4), a late-onset sensory and cerebellar ataxia, escaped detection for 25 years. Using long-read PacBio-HiFi and ONT-Nanopre sequencing and bioinformatic analysis, we identified expansion of a GGC DNA repeat in a >85% GC-rich region in exon 10 of the *ZFHX3* gene coding for poly-glycine (polyG). In a total of 15 nuclear families from Utah and 9 from Europe, the repeat was expanded to >40 repeats in SCA4 patients accompanied by significant phenotypic variation independent of repeat size compared to the most common normal repeat size of 21 repeats. The RE event likely occurred in a frequent Swedish haplotype shared by cases from Utah and Germany. Six characteristic ultra-rare SNVs in the vicinity of the RE in cases from Utah and Lübeck (Germany) indicate a common founder event for some of the patients. In fibroblast and iPS cells, the GGC expansion leads to increased ZFHX3 protein levels, polyG aggregates, and abnormal autophagy, which normalized with *ZFHX3* siRNA. Increasing autophagic flux may provide a therapeutic avenue for this novel polyG disease.

## Main Text

Cerebellar neurodegeneration has a prevalence exceeding that of motor neuron disease^1^. Remarkable progress has been made in the understanding of degenerative ataxias based on the identification of mendelian disease genes with the number of autosomal dominant ataxia genes now approaching 50, but a significant portion of familial ataxias has remained unidentified^2^. SCA4, an autosomal dominant cerebellar ataxia, was linked to chromosome 16q in a single pedigree in the US state of Utah^3^.

Using ancestry records of the Church of Jesus Christ of Latter-Day Saints, we were able to trace 15 nuclear families back to a likely common ancestor born in Southern Sweden at the turn of the 18^th^ and 19^th^ centurey (Supplemental Fig. 1). In contrast to other SCAs, the phenotype in these Utah pedigrees was characterized by prominent involvement of sensory nerves and neurons with relatively mild cerebellar involvement. A pedigree with a similar phenotype and also linking to chromosome 16q was subsequently identified in Germany with the exclusion of likely candidate genes containing DNA CAG repeats^4^.

Despite limiting the candidate region by genetic linkage analysis to a region of ∼6 Mbp, we were not able to identify likely pathogenic single nucleotide variants, indels or DNA repeat expansions, by conventional short-read next-generation sequencing using whole exome and whole genome technologies. This region of the human genome is GC-rich and contains a number of duplications and pseudogenes^5^.

To overcome these challenges we employed PacBio-HiFi and ONT-Nanopore sequencing technology that allows to capture long DNA reads from single strands of DNA. In order to detect the molecular cause underlying SCA4, comprehensive genetic studies were conducted with subsequent combined bioinformatic analyses of short- / long-read genome (SR-/LR-GS) and RNA-seq datasets. Variant filtering was done under the assumption of an autosomal dominant model of inheritance with a focus on rare variation in the linkage interval that might be challenging to detect or interpret (see Methods). First, SR-GS and RNA-seq data were analyzed according to established diagnostic standards using a pipeline optimized for variant detection ‘beyond the exome’ ^6^. This approach failed to detect any rare variation of likely clinical relevance.

Second, the generated LR-GS were searched for genomic variation within the linkage interval that might have been missed by previous variant calling algorithms. We identified 4 structural variants that were present only in affected family members, with only one of them, a GGC repeat expansion within exon 10 of *ZFHX3*, being rare and missed by previous variant calling algorithms. The structural variant was characterized as a 155 bp repeat expansion by TRGT^7^ and confirmed by independent discovery in the ONT sequencing data of the same samples. The repeat expansion was also found independently in two affected cases in Lübeck (Germany), again using HiFi sequencing and repeat expansion detection with TRGT. In order to provide further evidence of a disease-causal association of the *ZFHX3*-expansion with ataxia and sensory neuronopathy, we performed a targeted bioinformatic screen of an in-house database with 6,495 diagnostic-grade SR-GS datasets using ExpansionHunter (see Methods). In addition to successfully re-identifying the affected individuals of the Utah pedigree, this approach led us to detect expanded alleles in an additional 5 previously unsolved individuals with ataxia (Fig. 1).

**Fig. 1:**
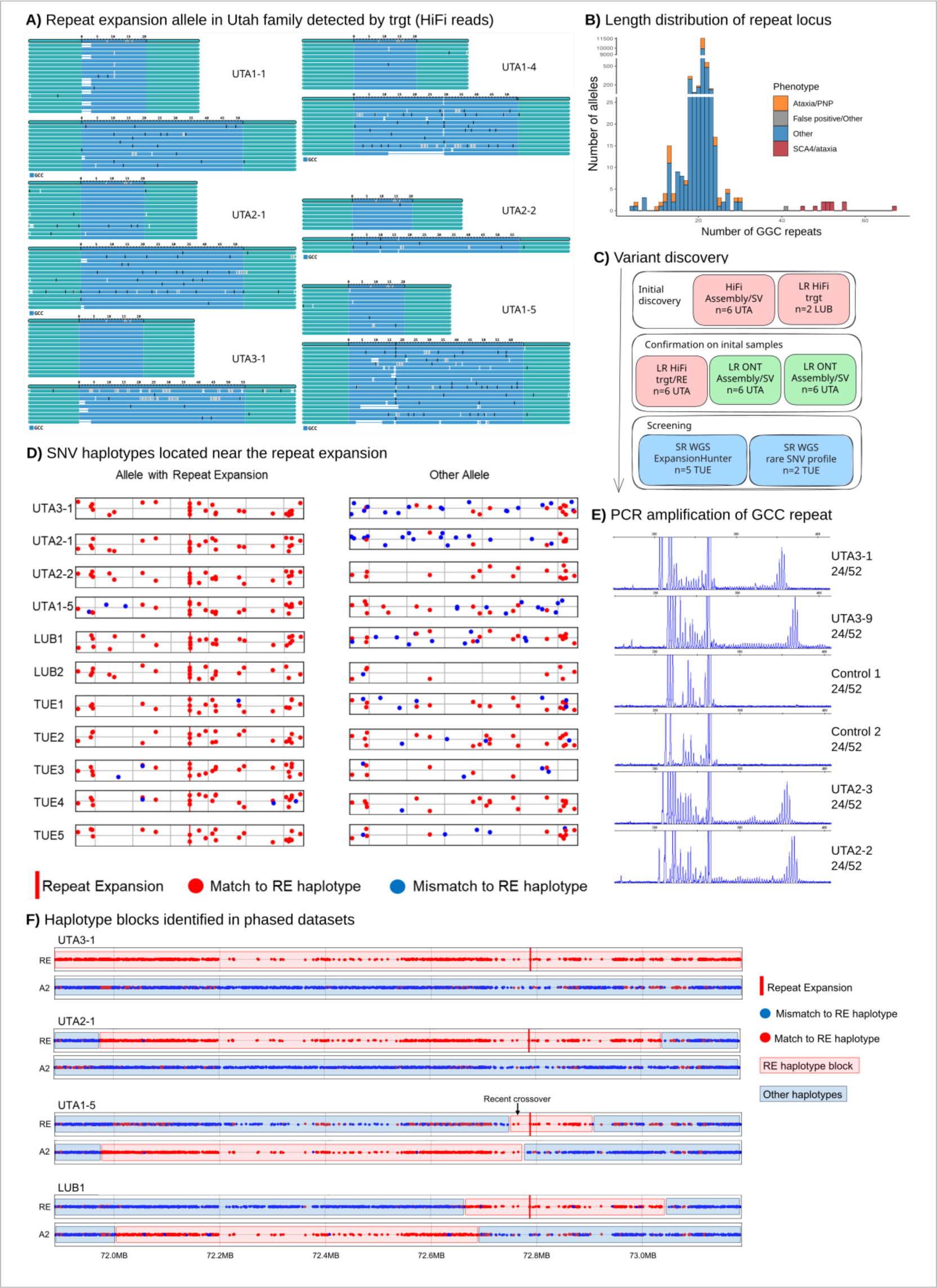
**Identification of the repeat expansion in *ZFHX3* as causative of SCA4**. Discovery of causal repeat expansion in Utah families 1-3. **A)** PacBio long-read sequencing data was used to identify the heterozygote allele expansions using TRGT. **B)** Screening of 6495 whole genome short read datasets (U. Tübingen) with ExpansionHunter revealed five additional cases as well as the length distribution of the GCC repeat. **C)** The repeat expansion was discovered by SV calling from *de novo* assembly and then confirmed using other long read approaches. Further screening of an in-house database revealed hits in 5 short read genomes. The RE in two additional HiFi samples were found independently in Lübeck. **D)** Close inspection of the SNVs in the RE-haplotype in the vicinity of the RE (+/- 50 kb) showed strong similarities between cases from Utah, Lübeck (Germany) and Tübingen (Germany). However, only the cases from Utah and Lübeck shared all 6 characteristic rare SNVs that reliably distinguish the RE-allele from other haplotypes (see Suppl. Table 2). **E)** PCR amplification of the GGC repeat using fibroblast cDNA from control and SCA4 individuals of the Utah pedigree. **F)** Haplotype phasing with long reads and comparison of phased haplotypes using one member of each Utah family and one case from Lübeck (Germany), revealed a large identical haplotype block (red bar). The RE-allele is always shown first. Cases UTA1-5 and LUB1 had recent crossover events close to the repeat expansion. Interestingly, we also found this RE-haplotype in a version without RE in the unaffected cases UTA2-2 and UTA2-4 (see Suppl. Fig. 2).

Long-read sequencing allows reliable haplotype phasing across long distances and is therefore highly suitable for disease haplotype analysis. We compared phased variants from one member of each Utah family and one Lübeck patient, each two of these individuals are separated by at least 6 generations (Suppl. Fig. 1). We defined the haplotype-phase of patient UTA3-1 containing the RE as template and identified a roughly 1Mb large haplotype shared by all four individuals, although with recent crossover-events in patients UTA1-5 and LUB1. RE discovery is summarized in Supplementary Table 1.

Interestingly, we find a highly similar haplotype in the unaffected cases UTA1-2 and UTA1-4 from Utah, showing only slightly increased differences in SNV content and, obviously, lacking the repeat expansion. We concluded that the repeat expansion event (RE) happened in a frequent Northern European haplotype, which exists in the same family (UTA1) in a RE and a non-RE version. To further define the haplotype, we screened for ultra-rare SNVs in the vicinity of the RE that distinguishes the RE and non-RE versions of the haplotype and found 6 SNVs within 75 kb of the RE unique to the alleles affected by the RE (Supp. Table 2). Four of these SNVs are directly next to the RE and expand the RE by generating new GGC triplets. Two SNVs are found upstream of the RE. Searching for this characteristic SNV profile led to the discovery of two additional cases in the in-house database previously missed by the ExpansionHunter screening.

Next, we plotted the SNV-similarity between all identified cases (4 from Utah, 7 from Germany) to ascertain, if a single founder event could explain all patients, or if independent founder events occurred. We found that SNV-similarities between the alleles affected by the RE are high (Fig. 1D), indicating a shared founder event.

However, only the cases from Utah and Lübeck share all 6 ultra-rare characteristic SNVs, while the five German cases sequenced by srWGS in Tübingen are missing one of these SNVs (Suppl. Table 2). This leads us to the most likely explanation that the RE haplotypes can be traced to a single founder. We assume that a single *de novo* SNV occurred after the repeat expansion event resulting in two distinguishable alleles.

PCR amplification of the expanded *ZFHX3*-GGC repeat proved to be extremely challenging using genomic DNA as a template owing to the extreme GC-richness (>85%) of exon 10 and flanking regions. Using cDNA from FBs we were able to amplify the normal and mutant repeat reliably using a highly customized PCR protocol (see methods section). Examples of PCR amplicons from control and SCA4 cDNAs are shown in Fig. 1F. As is typical for short tandem repeats, a pattern of shadow bands was detected and the tallest peak was assigned as the repeat for the respective individual. The repeat lengths detected by PCR were identical or within 2 repeat units with those assigned by long-read sequence analysis.

The ZFHX3 protein (also designated ATBF1) contains 3,703 amino acids (with 21 glycine residues) with a predicted molecular weight of 404 kDa. It is a transcription factor with functions as a tumor suppressor gene and as a risk allele for atrial fibrillation (AF)^8^. *ZFHX3* has abundant expression including in the nervous system^9^. *ZFHX3* loss-of-function mutations lead to a neurodevelopmental phenotype in humans associated with intellectual disabilities and facial dysmorphology^8^. Chromatin immunoprecipitation (ChIP) sequencing of human neural stem cells identified binding of *ZFHX3* to promoter regions, especially those implicated in expression regulation of genes in the Hippo/YAP and mTor pathways^8^.

With the discovery of the GGC expansion in *ZFHX3*, SCA4 can now be included in the small but growing group of disorders including fragile X-associated tremor/ataxia syndrome (FXTAS), neuronal intranuclear inclusion disease (NIID), and oculophryngodistal myopathy dystrophy (OPMD)^10–12^. Important genomic characteristics, however, set SCA4 apart from these disorders including size of the expanded GGC repeat, its location in a coding exon as compared to an untranslated region of the respective disease gene, and the function of the SCA4 gene as a transcription factor.

Table 1 shows the phenotypes of 3 families, of the several families identified, harboring polyG-expanded ZFHX3. In contrast to individuals harboring loss-of-function *ZFHX3* mutations, SCA4 patients show no neurodevelopmental orb dysmorphology phenotypes. Adult-onset cerebellar and sensory ataxia was present in all families; gait ataxia was the presenting sign in almost all families. Some individuals had additional symptoms of dysphagia and autonomic dysfunction. Similar to what has been observed in individuals with SCA27b^13,14^, some individuals had chronic cough without obvious cause after intensive investigation.

**Table 1:** SCA4 phenotype. Note that n/a indicates not applicable or no data.

|  | SCA4 Utah PMID: 8755926 | SCA4 Lübeck PMID: 12796826 | München (Family) |
| --- | --- | --- | --- |
| **published** | published 1996 | published 2006 | no |
| **Family members** | 58 verified affected family members, 526 7-generation family | 31 affected family members, 6 generations | 6 affected family members, 4 generations |
| **Age of onset** | 12-57 (median 34 years) | 20-61 year (median 38.3 years) | 20-61 year (median 38.3 years) |
| **First symptom** | gait disturbance | gait ataxia and dysarthria | gait ataxia and dysarthria |
| **Ataxia** | all | all | all |
| **Ocularmotor function** | n/a | saccadic pursuit | n/a |
| **Dysarthria** | 50% | all | all |
| **Neuropathy (impaired vibration)** | 95% had vibratory and joint-position sense loss | reduced | n/a |
| **Nerve conduction** | 13 patients - sensory and/or motor neuropathy | absent sural sensory nerve action potential | n/a |
| **Cough** | 40% | chronic cough | chronic cough |
| **Dysphagia** | 60% | n/a | n/a |
| **Cognition and behavior** | 1 affected female dx as bipolar | n/a | n/a |
| **Dysautonomia** | neurogenic orthostatic hypotension | neurogenic orthostatic hypotension | n/a |
| **MRI** | n/a | cerebellar atrophy | n/a |
| **Other** | exercise induced dystonia, tongue fasciculations, tremor | Babinski sign, limb dysmetria | n/a |

Signs of cerebellar dysfunction were reflected in loss of volume in specific CNS structures. Representative MRI imaging of an SCA4 patient 10 years after disease onset is shown in Fig. 2A-C. There is relatively mild cerebellar atrophy without the middle-cerebellar-peduncle sign typical for Fragile X-associated tremor/ataxia syndrome (FXTAS)^15^ or the corticomedullary junction white matter changes typically seen in nuclear intranuclear inclusion disease (NIID)^16,17^. On the mid-sagittal T1-weighted MRI, midline cerebellar atrophy and minimal pontine atrophy can be appreciated with an upper cervical cord of reduced diameter (Fig. 2C).

**Fig. 2:**
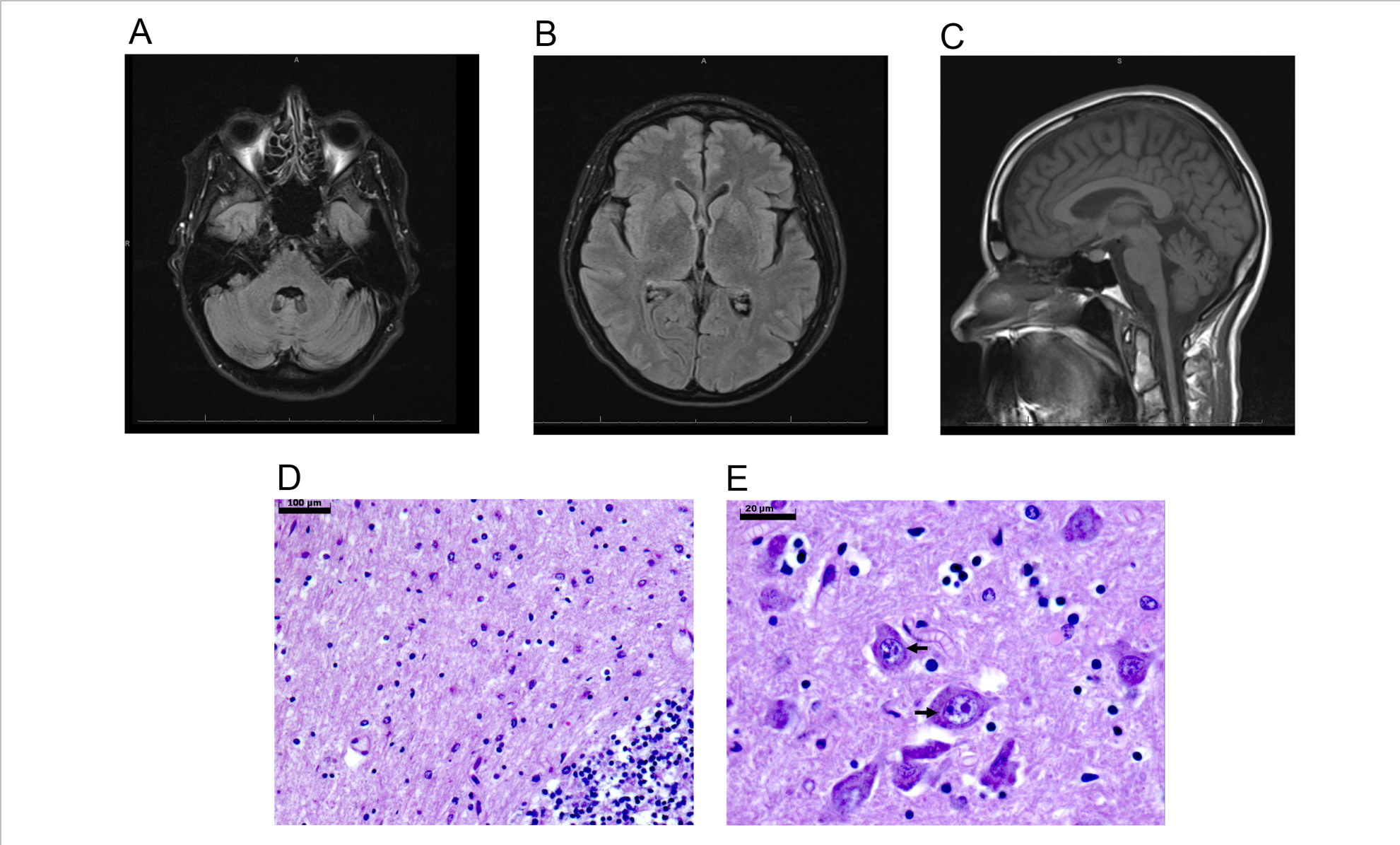
MRI and histo-pathological features of SCA4 patients. (A-C) MRI imaging of an SCA4 patient 10 years after symptom onset. **A)** Axial FLAIR MRI of the posterior fossa with absent hyperintensities in the middle cerebellar peduncles **B)** Axial FLAIR MRI of the cerebral hemispheres, note lack of cortico-medullary hyperintensities. **C)** Mid-sagittal T1-weighted MRI showing mild cerebellar and spinal cord atrophy. **D)** Hematoxylin/eosin stained section of the cerebellar vermis of an individual with long-duration SCA4 shwoing absence of Purkinje cells and rarefaction of granule cells. **E)** Hematoxylin/eosin stained section of the base of the pons showing amphophilic intranuclear neuronal inclusions (arrows). Bar: 100 μm (D), 20 μm (E).

Hematoxylin/eosin-stained slides were available from an autopsy of a brain of a male in his mid-70s who died >30 years after onset of symptoms of gait ataxia (Fig. 2D,E). The cerebellum was largely depleted of Purkinje cells with a reduction in the width of the molecular layer. The dentate nucleus was spared. In the base of the pons, several neurons contained intranuclear inclusions. The inclusions were distinct from nucleoli by their slightly larger size and amphophilic staining. These findings are similar as one prior descriptions of an SCA4 brain^18^, except for the lack of inclusions in the latter study.

The location of the GGC repeat in a predicted coding exon of *ZFHX3* suggested that the repeat was translated to a polyG domain that was longer than in controls and in-frame with the rest of the ZFHX3 protein. We used an antibody to the ZFHX3 protein and a monoclonal antibody to polyG repeats to analyze expression in protein extracts from cultured fibroblasts (FBs) derived from four individuals with SCA4 and controls (Fig. 3). The polyclonal ZFHX3 antibody recognized a ∼400 kD band consistent with the full-length ZFHX3 protein. To determine the specificity of this band, we incubated HEK-293 cells for 72 hrs with 2 different concentrations of a validated siRNA to *ZFHX3*.

**Fig. 3:**
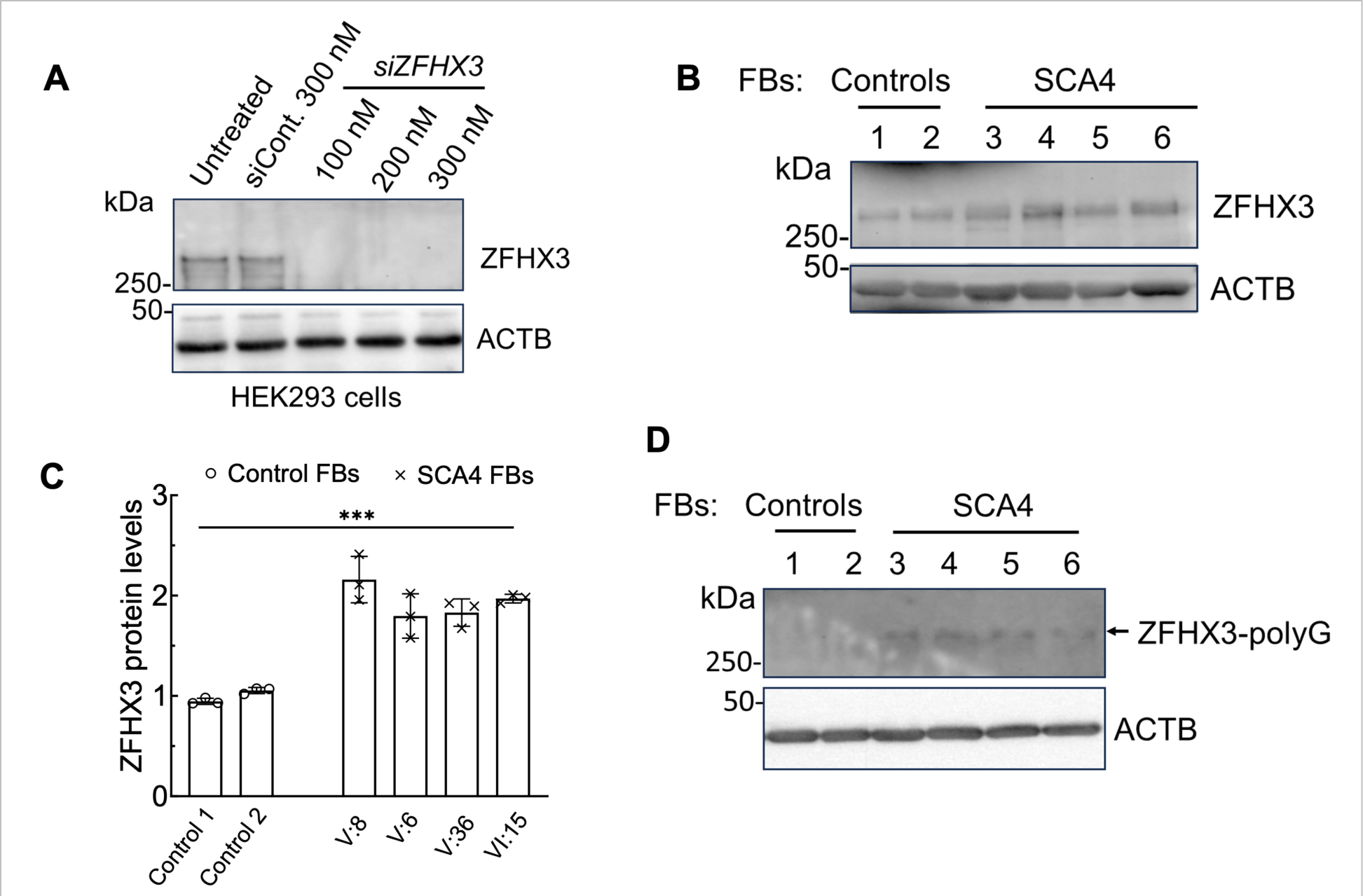
SCA4 patient FBs show ZFHX3 expression. (A) Characterization of the ZFHX3 antibody. The ZFHX3 antibody demonstrates specificity for the detection of endogenous human ZFHX3. HEK-293 cells were transfected with control and human *ZFHX3* siRNAs at indicated concentrations and harvested at 3 days post-transfection. Protein extracts were analyzed by western blotting to verify ZFHX3 silencing. **(B)** Western blot analysis of FB protein extracts from two controls (lanes 1 and 2) and four SCA4 patients (V:8, V:6, V:37, and VI:18; lanes 3,4, 5, and 6). The patient FBs show increased ZFHX3 levels. **(C)** ZFHX3 quantification relative to ACTB of three independent experiments. Two-way ANOVA followed by Bonferroni tests. Data are mean ± SD, ***p < 0.001. **(D)** Anti-polyG antibody detects mutant ZFHX3 protein in all four SCA4 FBs, but not in control FBs on western blot. ACTB was used as a loading control.

Treatment with the *ZFHX3* siRNA, but not with a control siRNA, resulted in disappearance of the 400 kDa band (Fig. 3A). This established that the 400 kDa protein was indeed encoded by *ZFHX3*.

We next examined whether the mutant protein was altered in its overall abundance and found that SCA4 protein sample had increased ZFHX3 protein, although levels showed some variation among the different lines (Fig. 3B,C). These results are consistent with a gain-of-function of polyG-expanded ZFHX3, but at this point we cannot discriminate between a gain-of-normal as compared to a gain-of-toxic function.

We then tested whether the ZFHX3 protein expressed in SCA4 FBs contained a polyG domain. To do this, we used a monoclonal antibody that recognizes expanded polyG domains^19^. This antibody recognized a 400 kDa protein in fibroblast cells from 4 SCA4 individuals, but did not recognize a signal in control FBs (Fig. 3D). It is likely that this antibody recognizes a specific conformation in expanded polyG domains similar to the 1C2 antibody for polyglutamine domains^20^.

The presence of inclusions in SCA4 brain sections and elevated ZFHX3 in SCA4 FBs prompted us to evaluate ZFHX3 abundance and aggregation in cultured cells. However, with extant antibodies to ZFHX3 or polyG we were not able to detect these in FBs. We therefore produced SCA4 patient derived iPSCs with the goal of generating iPSC- derived neurons. Whereas control iPSCs were easily differentiated into neurons using an established protocol, SCA4 iPSCs became rapidly apoptotic upon induction of differentiation. Thus, we were limited to analyze ZFHX3 in iPSCs. When we assayed ZFHX3 abundance in SCA4 iPSCs at various passages, we detected a significant increase in abundance of the protein by western blot analysis (Fig. 4A,B). Although polyG aggregates were not discernible in SCA4 FB cell lines, an abundance of SCA4 iPSCs contained polyG aggregates that also labeled positively for ubiquitin (Fig, 4C-F). The aggregates were usually multiple and large. Their location was clearly cytoplasmic and no intranuclear aggregates were detected. These findings are reminiscent of mislocalisation seen with mutant TDP-43, an RNA/DNA binding protein, leading to pathogenetic hypotheses incorporating loss of nuclear function and cytoplasmic gain-of- toxic function via aggregate formation^21^. Further studies are needed to determine whether pathogenesis of polyG mutations in ZFHX3 shares features of protein mislocalization known for TDP-43.

**Fig. 4.**
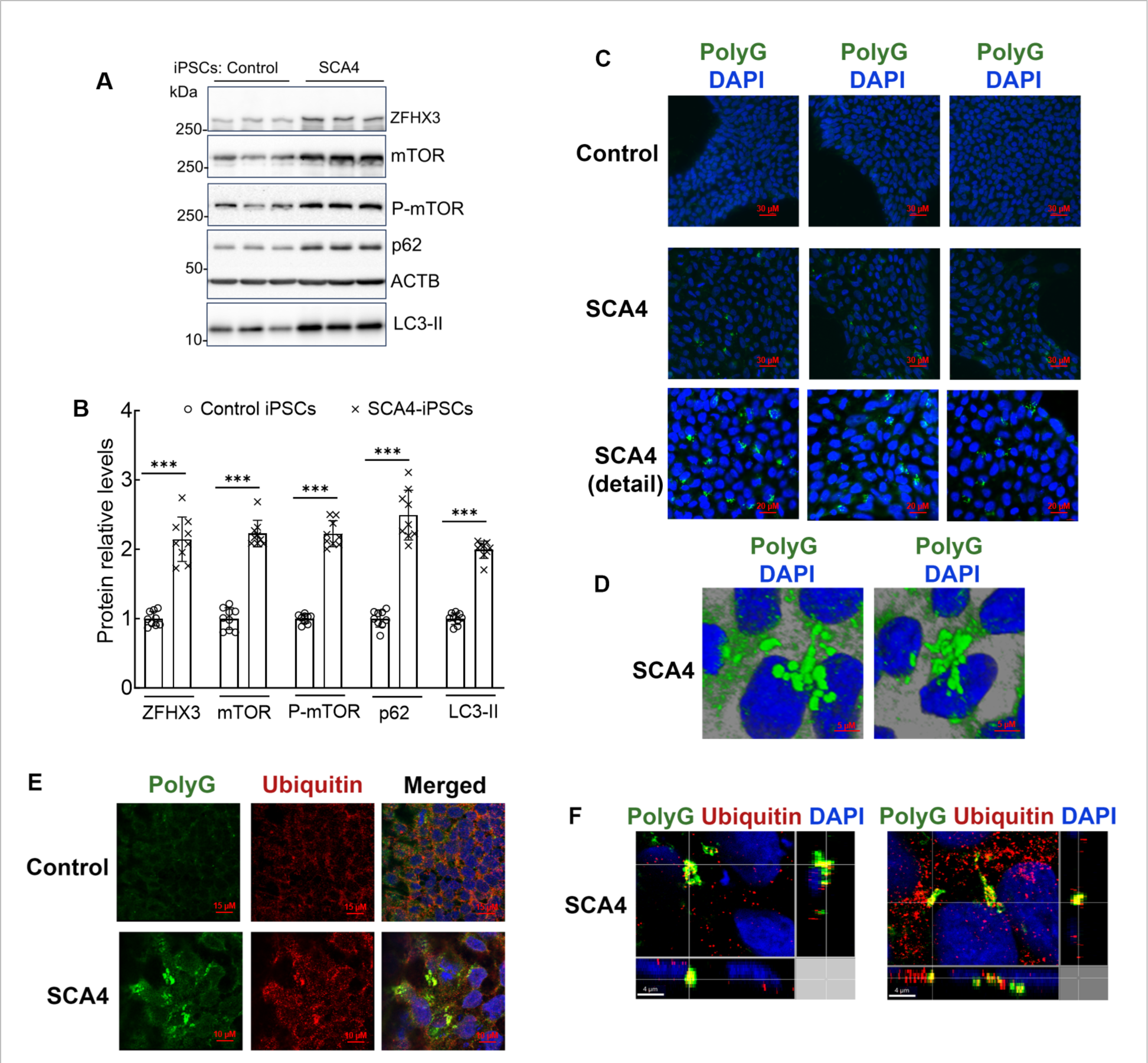
SCA4 patient-derived iPSCs display disease phenotypes. (A) Western blot analysis of protein extracts from iPSCs of SCA4 (V:8) iPSCs show increased levels of ZFHX3, mTOR, P-mTOR, p62, and LC3-II compared to the normal control. ZFHX3 protein was detected with a ZFHX3 polyclonal antibody (ThermoFisher, USA, Cat# PA5-118401). Each lane is an independent cell pellet harvested at a different passage. Panels correspond to representative images of 3 independent western blots. **(B)** Protein levels were normalized to ACTB, and quantification of average fold changes for indicated proteins are shown. Two- way ANOVA followed by Bonferroni tests. Data are mean ± SD, ***p < 0.001. **(C)** Immunofluorescence of control and SCA4 iPSC show poly-G aggregates (green) and nuclei (blue) **(D)** Tridimensional rendering of polyG aggregates and nuclei of SCA4 iPSC. **(E)** Immunofluorescence for polyG (green) and Ubiquitin (red), show an overall view in control iPSC and a zoomed in view in SCA4 iPSC. (F) Confocal microscopy sectioning of SCA4 iPSC in lateral views indicate partial colocalization of polyG and Ubiquitin signals (yellow).

We had previously shown that autophagic flux was impaired in cellular and animal models expressing mutant TDP-43 or mutant ATXN2 ^22–24^. SCA2 is caused by expansion of a polyQ domain in ATXN2 ^25^ and is also associated with cytoplasmic aggregates^26^. We therefore examined whether cellular models of SCA4 showed a similar alteration of autophagy (Fig. 4 A,B). We determined abundance of phosphorylated (active)- and total mTOR in protein extracts. We also measured p62 and LC3-II, both of which increase in abundance with reduced autophagy^27^ (Fig. 4A). In triplicate experiments, we found that all 4 proteins had increased steady-state levels in SCA4 iPSCs consistent with abnormal autophagy (Fig. 4C).

With the availability of a larger number of FB lines from different individuals, we characterized autophagy in this cell type. As in iPSCs, we detected an increase in mTOR, p-mTOR, and p62 (Fig. 5 A,B). Wildtype ATXN2 was significantly upregulated as well suggesting it as a potential target for SCA4 therapy using ASOs to *ATXN2*. Of note, concurrent pathologic CAG-repeat expansion in *ATXN2* was excluded in all cell lines.

**Fig. 5:**
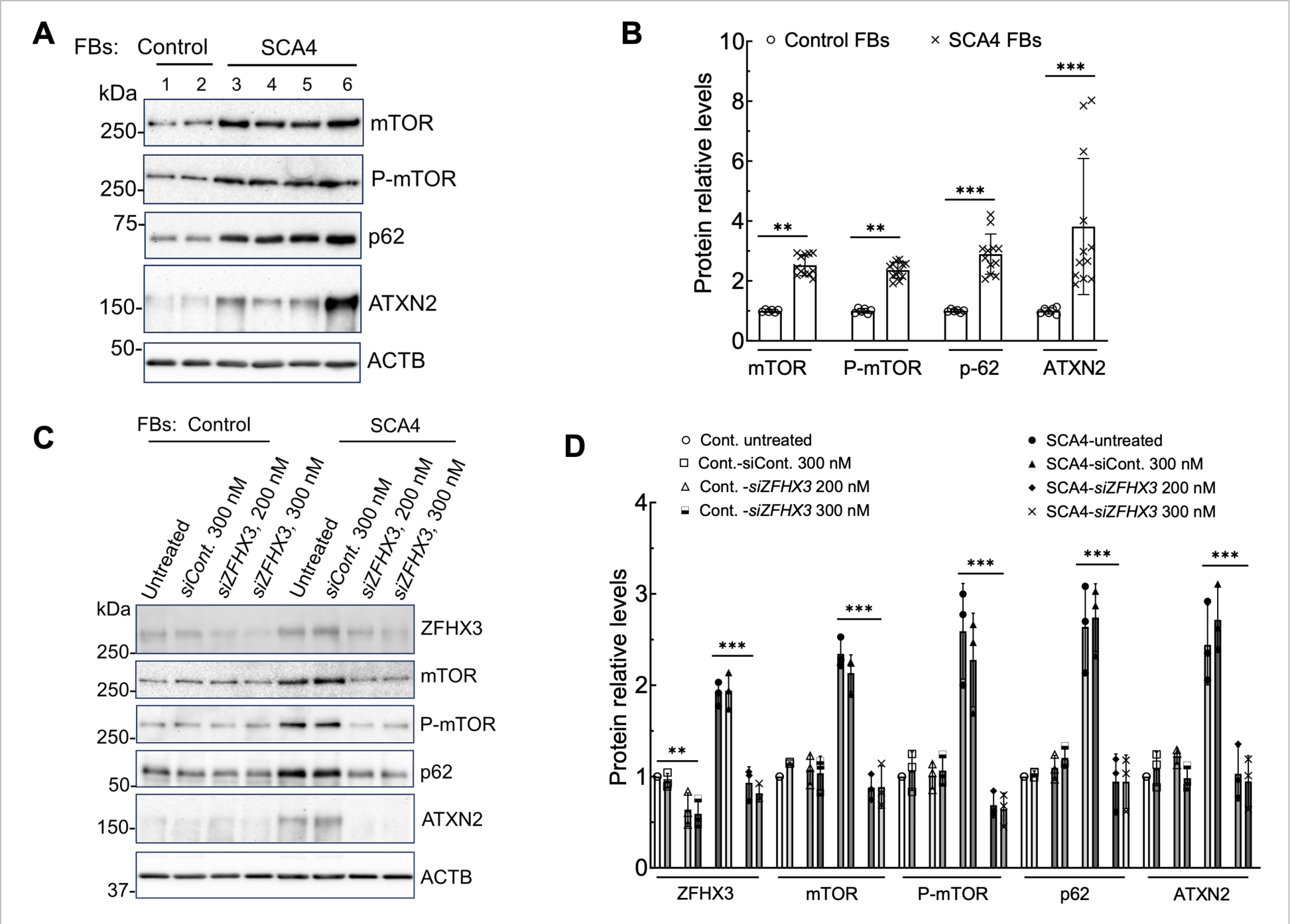
SCA4 FBs show increased ATXN2 levels and abnormal mTOR signaling. A) Western blot analysis of protein extracts from two controls (1 and 2; lanes 1 and 2) and four SCA4 patient-derived FBs (V:8, V:6, V:37, VI:18; lanes 3,4, 5, and 6) show increased levels of mTOR, P-mTOR, p62 and ATXN2 compared to normal control FBs. (B) Quantifications relative to ACTB. Representative blots of 3 independent experiments are shown. (C, D) RNAi-mediated *ZFHX3* depletion in SCA4 patient FBs normalized autophagy pathway protein levels. (C) Control (#1) and SCA4 (V:8) FBs were treated with siRNA to *ZFHX3* for 4 days and analyzed by Western blotting. Reducing expression of ZFHX3 results in lowered mTOR, P-mTOR, p62, and ATXN2 levels in SCA4 FBs. ZFHX3 protein was detected with anti-ATBF1/ZFHX3 (AT-6) polyclonal antibody (MBL International Corporation, Cat# PD011] on western blot. ACTB is used as a loading control, and representative blots of 3 independent experiments are shown. (D) Quantifications relative to ACTB. Ordinary one-way ANOVA followed by Bonferroni tests for multiple comparisons. Data are mean ± SD, ns = p > 0.05, ***p < 0.001.

To determine whether altered autophagy was directly related to expression of endogenous mutant *ZFHX3*, we used RNA interference (RNAi) to perform knockdown (Fig. 5C, D). Reduction of ZFHX3 normalized all autophagic markers and reduced ATXN2 levels. These results are consistent with a major role in pathological autophagy regulation by mutant ZFHX3, but do not exclude other pathomechanisms such as nuclear RNA toxicity or RAN-translation. Intriguingly, they also suggest crosstalk between the polyQ protein ATXN2 and ZFHX3 polyG pathology. Reduction of ATXN2 by treatment with ASOs with the MOE gapmer chemistry^28^ improved neurodegeneration in mouse models of SCA2 and ALS^29,30^; an ASO to *ATXN2* is currently in a phase 1 trial for sporadic ALS (NCT04494256). Future studies will need to address whether reduction of wildtype ATXN2 can reduce ZFHX3-polyG toxicity.

In summary, we identified a dominant GGC repeat expansion in *ZFHX3*. Our study underscores the importance of identifying genetic variation including novel repeat expansions in extremely GC-rich genomic regions, which may account for some of the missing heritability in neurodegenerative disorders as mendelian or risk alleles. Our findings add to the list of neurodegenerative diseases caused by poly-G expansions^11^, which can also manifest by repeat associated non (RAN) -AUG translation of non-coding regions as in *NOTCH2NLC*^31^ and *FMR1*^32,33^. SCA4 differs phenotypically from other polyG disorders in the absence of episodic phenomena, myopathy, and white matter changes and presence of cytoplasmic polyG-inclusions, impaired autophagy and expression of the repeat at the C-terminus of a transcription factor^34–38^.

## Methods

### Human subjects

All procedures involving human subjects were approved by the Institutional Review Board (IRB) at the University of Utah. Human subjects use was approved by the ethics committee of the Medical Faculty of the University of Tübingen, Germany (Genome+, ClinicalTrial.gov-Nr: NCT04315727). All human subjects in the US and Europe gave written consent for inclusion in the study.

### Short-read genome sequencing and analysis

Genome analyses were performed on six affected and two healthy individuals from family 1 at the Institute of Medical Genetics and Applied Genomics (IMGAG), University Hospital Tübingen, Germany. Genomic DNA was extracted from whole blood using the FlexiGene DNA kit (Qiagen, Hilden, Germany) and quantified using the Qubit Fluorometer (Thermo Fisher Scientific, Dreieich, Germany). One µg of genomic DNA was further processed using the TruSeq PCR-Free Library Prep kit (Illumina, Berlin, Germany) and generated libraries were sequenced on a NovaSeq6000 System (Illumina) as 2x150 bp paired-end reads to an average 49X coverage.

Mapping, variant calling and annotation of the data was performed using the megSAP pipeline (https://github.com/imgag/megSAP) developed at the Institute of Medical Genetics and Applied Genomics, University Hospital Tübingen, Germany. Details about the used tools and databases can be found in the megSAP documentation and in our previous publication^6^.

Variant filtering and interpretation include various filtering steps to prioritize potentially clinically relevant variants with a focus on alterations in the linkage interval. In a stringent multi-sample analysis, we searched for potentially protein-altering variants with a minor allele frequency (MAF) <0.1% both in gnomAD^39^ and an in-house database (>20,000 ES and GS datasets from unrelated phenotypes). This stringent filtering failed to identify any potentially causal variant and was subsequently expanded to include less rare (MAF < 1%) SNVs and Indels in the coding and non-coding regions of the linkage interval as well as copy number alterations and complex structural variants. For none of the detected variants any effect was observed on RNA expression levels and the OMIM-listed genes in neighboring coding regions have not been associated with overlapping phenotypes.

### Long-read HiFi sequencing

HiFi whole genome sequencing was performed for six affected and two healthy individuals from the Utah pedigree. Genomic integrity was assessed using pulse-field capillary electrophoresis with the Genomic DNA 165 kb Analysis Kit on a FemtoPulse (Agilent) instrument. Quantitation of DNA was assessed using the dsDNA High Sensitivity assay on a Qubit 3 fluorometer (Thermo Fisher). A total of 5 µg of genomic DNA was sheared with Megaruptor 3 (Diagenode) . Libraries were prepared with the HiFi SMRTbell Library Preparation Kit TPK 2.0 (Pacific Biosciences). Size fractionation of SMRTbell libraries was prepared with the BluePippin System (Sage Science) for removal of libraries with <10kb in size. SMRTbell libraries were assessed with the Genomic DNA 165 kb Analysis Kit on a FemtoPulse (Agilent) instrument. SMRTbell libraries were prepared with the Sequel II binding kit 2.2 (Pacific Biosciences) and sequenced with SMRTCell 8M with a Sequel II instrument (Pacific Biosciences) at a loading concentration of 70-90 pM. The mean HiFi read length was 16 kb. The average coverage was 31.

HiFi reads were assembled using Hifiasm^40^ after quality control with genomescope^41^. We then used the hapdup-hapdiff pipeline^42^ to create haplotype phased assemblies and call structural variants between the assembly and the GRCh38 reference genome. Small variants were called with Deepvariant^43^. Structural variants were merged with SURVIVOR^44^ to reveal mutations present only in the affected samples. Finally, characterization of the repeat expansion was done using TRGT^7^ with a slightly modified version of the full repeat catalog that contains extended coordinates for the *ZFHX3* repeat.

### Long-read ONT sequencing

ONT whole genome sequencing was performed for six affected and two healthy individuals from the Utah pedigree. Genomic integrity was assessed using pulse-field capillary electrophoresis with the Genomic DNA 165 kb Analysis Kit on a FemtoPulse (Agilent) instrument. Quantitation of DNA was assessed using the dsDNA High Sensitivity assay on a Qubit 3 fluorometer (Thermo Fisher) and purity was assessed by Nanodrop. A total of 3 µg of genomic DNA was sheared with Megaruptor 3 (Diagenode) for samples harboring high-molecular weight DNA fragments. Libraries were prepared with the 1D Ligation SQK-LSK109-XL Sequencing kit (Oxford Nanopore Technologies). A total of 600 ng (50 fmol) of each library was loaded on a single PromethION R9 flow cell. A nuclease flush was performed to re-load 350-600 ng of library when possible.

The average coverage was 27 reads. Long-read structural variant analysis

A *de novo* genome assembly using Nanopore reads was generated with flye^45^. We resolved individual haplotypes and phased structural variants from the assemblies using hapdup-hapdiff. Additionally, structural variants were also called with Sniffles2 ^46^. In a second, alignment based approach, the long reads were mapped to the human reference genome GRCh38 using minimap2 ^47^. Structural variants were called using Sniffles2 ^46^, small variants were called using Pepper-Margin-Deepvariant^48^. All steps of the ONT analysis were done using the megLR pipeline available on GitHub.

Long reads were mapped to the human reference genome GRCh38 using minimap2 ^47^. The average coverage was 27 reads overlapping at least one flanking region of the RFC1 repeat location (chr4:39348424-39348479) were analyzed for repeat length and motif. Analysis scripts are publicly available (https://github.com/caspargross/expander Commit #37687e3).

### RNA-seq and expression analysis

RNA-seq was performed on ten cases. RNA was extracted from cultivated fibroblasts with QIAsymphony RNA kits on a QIAsymphony SP with the protocol RNA CT 400 V7. From 100 ng of total RNA, mRNAs were enriched using polyA capture on a NEBNext Poly(A) mRNA Magnetic Isolation Module (NEB). Libraries were prepared on a Biomek i7 (Beckman Sequencing) using Next Ultra II Directional RNA Library Prep Kits for Illumina (NEB) according to the manufacturer’s instructions. The fragment sizes were determined with a Fragment Analyzer (High NGS Fragment 1-6000bp assay (Agilent)) and the library concentration (approximately 5 ng/µl) was analyzed with an Infinite 200Pro (Tecan) and the Quant-iT HS Assay Kit (Thermo Fisher Scientific). 215 pM cDNA libraries were sequenced as 2x100 bp paired-end reads on an Illumina NovaSeq6000 (Illumina, San Diego, CA, USA) with approximately 50 million clusters per sample.

Generated RNA sequences were analyzed with respect to aberrant expression, aberrant splicing, and allelic imbalance using the megSAP pipeline (version 2022_08, https://github.com/imgag/megSAP). In brief, the ngs-bits tool collection (version 2022_08-92, https://github.com/imgag/ngs-bits) was used for quality control (ReadQC) and pre-processing (SeqPurge) of fastq files. STAR (version 2.7.10a, https://www.ncbi.nlm.nih.gov/pubmed/23104886, https://github.com/alexdobin/STAR/) was used for read alignment and detection of splice junctions, which were postprocessed with SplicingToBed. After mapping, MappingQC was used for quality control and Subread (version 2.0.3, https://pubmed.ncbi.nlm.nih.gov/30783653/, https://sourceforge.net/projects/subread/) for read counting based on an Ensembl gene annotation file (GRCh38, release 107, http://www.ensembl.org/index.html). Upon normalization (megSAP) and quality assessment (RnaQC), expression values of genes and exons were compared with an in-house cohort (same tissue and processing system) using NGSDAnnotateRNA.

Clinical interpretation was done with GSvar, a in-house diagnostics software developed at IMGAG. GSvar allows filtering for expression of genes and exons by gene, biotype, expression value, read counts, and Z-score compared to the cohort and the splice junctions by gene, type, read count, and motif. Integrative Genomics Viewer (IGV, version 2.11.9, https://www.nature.com/articles/nbt.1754, https://software.broadinstitute.org/software/igv/) was used for visual inspection.

### Repeat screening of an in-house cohort

To determine the distribution of the *ZFHX3* repeat sizes in cases and controls, a screen of 6,495 genome datasets was performed using ExpansionHunter^49^. The following JSON repeat definition was used (coordinates are for GRCh38):

{

“LocusId”: “ZFHX3”,

“LocusStructure”: “(GCC)*”,

“ReferenceRegion”: “chr16:72787695-72787757”, “VariantType”: “Repeat”

}

Note that repeats designated “normal” encode 6 glycines interrupted by a single serine and are followed by a variable tract of approximately 14 glycines, and for example, a normal repeat length of 21 includes the serine residue (G6SG14). Expanded repeats are uninterrupted GCC repeats, at DNA level.

We also screened 15,281 exome datasets including positive controls, but no repeat expansion was identified. This could mean that the repeat expansion cannot be detected in exome datasets. We believe that the longer read length (150bp vs. 100pb) and bigger insert size (400bp vs. 200bp) of genomes are crucial to reliably identify expanded alleles.

### Reverse transcription-PCR (RT-PCR)

GGC repeat lengths in *ZFHX3* exon 10 were determined by RT-PCR. Reactions were 20 μl, including 4 μl 5X SuperFi II buffer, 0.4 μl 10 mM dNTPs (New England Biolabs, Cat# N0447), 2 μl 5 mM 7-deaza-dGTP (New England Biolabs, Cat# N0445), 4.2 μl nuclease-free water, 0.4 μl Platinum SuperFi II DNA Polymerase (ThermoFisher Scientific, Cat# 12361050), 2 μL of each 3 μM forward (FAM label) and reverse primer (0.3 μM) and 25 ng cDNA. Thermal cycling conditions were 98 °C for 5 min, 5 cycles of 98 °C for 1 min, 65 °C for 20 s, 72 °C for 2 min 30 s, 30 cycles of 98 °C for 1 min, 60 °C for 20 s, 72 °C for 2 min 30 s and a final extension at 72 °C for 5 min. PCR cycling was performed on a SimpliAmp Cycler (ThermoFisher Scientific, Cat# A24811). 2ul of the PCR amplicon is then loaded to instrument Applied Biosystems 3730xl DNA Analyzer, utilizing 50cm capillary, on POP7 polymer, and results were analyzed using the GeneMapper Software v3.7 (Applied Biosystems), using the GS 500 LIZ size standard (Applied Biosystems). Primers used were KPF-10F (Fam Labeled) 5’-TTTGGCGTTTCTTGCTGCTC-3’ and KPF-10R 5’-ACTCCCTCTACGACCCCTTC 3’.

The expected amplicon size was 356 bp for a fragment containing 21 repeats. Cell culture, and transfections Primary skin cell fibroblast (FB) cultures were established from two non-SCA4 control individuals and four SCA4 patients. These included 2 normal control lines (21/21), and 4 SCA4 lines: V:8 (21/46), V:6 (21/51), V:37 (22/48), VI:18 (21/53), where repeat lengths are in indicated parentheses. Human FBs were cultured and maintained in Dulbecco’s Modified Eagle’s (DMEM) medium containing 15% fetal bovine serum (FBS), as previously described^22^. Transfections of siRNAs were performed as previously described^22–24^. Briefly, cells were transfected with siRNAs using the Lipofectamine 2000 Transfection Reagent (ThermoFisher, USA, Cat# 11668019) according to the manufacturer’s protocol. Cells were harvested 4–5 days post-transfection.

### Induced pluripotent stem cells (iPSCs)

Human iPSC lines from control #1 and SCA4 line V:8 FB cultures were used for the production of iPSCs. iPSC lines were generated by transfection of a single mammalian episomal expression plasmid (pCEP4-4f) that we modified to express each of the four Yamanaka reprogramming factors (SOX2, OCT3/4, c-MYC and KLF4) each under the control of individual CMV promoters. This work closely followed published protocols^50–52^. Briefly, human control #1 and SCA4 line V:8 FBs were electroporated with the pCEP4- 4f reprogramming plasmid using the NeonTransfection System (ThermoFisher, Cat# MPK10025), then plated on vitronectin coated six well plates, and cultured with DMEM with 15% FBS for 5 days. Subsequently, the culture medium was changed to Essential 6 media (ThermoFisher Cat# A1516401) supplemented with basic fibroblast growth factor, bFGF (ThermoFisher, Cat# PHG0264) every other day until the emergence of iPSC-like colonies, which occurred approximately 23-25 days post-transfection. These iPSC colonies were then harvested and maintained in Essential 8™ Medium (ThermoFisher, Cat# A1517001) in vitronectin coated plates.

### siRNAs and reagents

The siRNAs used in this study are as follows: All Star Negative Control siRNA (Qiagen, Cat# 1027280), human siZFHX3: 5’-AGAAUAUCCUGCUAGUACAdTdT-3’ ^53,54^. All siRNA oligonucleotides were obtained from Integrated DNA Technologies, USA. Before use, the oligonucleotides were deprotected and the complementary strands were annealed.

### Preparation of protein lysates and Western blotting

Cellular extracts were prepared by a single-step lysis method as previously described^22–24^. The harvested cells were suspended in SDS-PAGE sample buffer (Laemmli sample buffer, Bio-Rad Cat #161–0737) and then boiled for 5 minutes. Equal amounts of the extracts were used for Western blot analyses. Protein extracts were resolved by SDS-PAGE and transferred to Hybond-P polyvinylidene difluoride (PVDF) membranes (Amersham Bioscience, Chicago, IL), and then processed for Western blotting according as previously described^22–24^. Immobilon Western Chemiluminescent HRP Substrate (EMD Millipore, Billerica, MA; Cat# WBKLSO500) was used to visualize the signals, which were detected on the ChemiDoc MP imager (Bio-Rad Laboratories). The band intensities were quantified by ImageJ software analyses after inversion of the images. Relative protein abundances were expressed as ratios to β-actin (ACTB) or glyceraldehyde-3-phosphate dehydrogenase (GAPDH).

### Antibodies used for Western blotting

The antibodies used for western blotting and their dilutions were as follows: mouse anti-Ataxin-2 antibody (Clone 22/Ataxin-2) [(1:4000), BD Biosciences, Cat# 611378]; LC3B Antibody [(1:7000), Novus biologicals, NB100-2220]; monoclonal anti-β- Actin−peroxidase antibody (clone AC-15) [(1:30,000), Sigma-Aldrich, A3854]; SQSTM1/p62 antibody [(1:4000), Cell Signaling, Cat# 5114]; mTOR antibody [(1:4000), Cell Signaling, Cat# 2972]; Phospho-mTOR (Ser2448) antibody [(1:3000), Cell Signaling, Cat# 2971]; and GAPDH (14C10) rabbit mAb [(1:7,000), Cell Signaling, Cat# 2118]. We used each of these antibodies in our previous publications^22–24^. Additional antibodies for western blotting were anti-polyG mouse monoclonal antibody, clone 9FM- 1B7 [(1: 3000), Sigma-Aldrich, Cat# MABN1788], ZFHX3 polyconal antibody [(1:3000), ThermoFisher, Cat# PA5-118401], and anti-ATBF1/ZFHX3 (AT-6) (Human) polyclonal antibody [(1:3000), MBL International Corporation, Cat# PD011]. The secondary antibodies were Peroxidase-conjugated AffiniPure goat anti-rabbit IgG (H + L) antibody [(1:5000), Jackson ImmunoResearch Laboratories, Cat# 111-035-144], and anti-mouse IgG, HRP-linked Antibody [(1:5000), Cell Signaling, Cat# 7076].

### RNA expression analyses by quantitative RT-PCR

Total RNA was extracted from harvested cells using the RNaeasy mini kit according to the manufacturer’s protocol (Qiagen, USA). DNAse I treated RNAs were used to synthesize cDNA using the High-Capacity cDNA Reverse Transcription Kit (ThermoFisher, Cat# 4368814). Quantitative RT-PCR was performed in QuantStudio 12K (Life Technologies, Inc., USA) at University of Utah core facilities. Taqman assays were performed using the following assay reagents: Human ZFHX3 [Assay ID: Hs00199344_m1 (Probe 1; Exons 5 and 6)]; Human ZFHX3 [Assay ID: Hs00994905_m1 (Probe 2; Exons 9 and 10)]; Human ACTB (Assay ID: Hs01060665_g1) (ThermoFisher Scientific, USA).

### Immunofluorescent labeling and antibodies used

iPSC were plated on glass coverslips on a Geltrex substrate (ThermoFisher Scientific, USA). Cells were fixed with 4% paraformaldehyde and blocked and permeabilized with 5% goat serum and 0.1% Triton X-100 (ThermoFisher Scientific, USA). The primary antibodies utilized were Anti-FMR1polyG Antibody (Millipore-Sigma MABN1788) and Anti-Ubiquitin (abcam ab134953), both at a dilution of 1:100 with overnight incubation. The secondary antibodies were Goat anti-Mouse Alexa Fluor Plus 488 or 594 (ThermoFisher Scientific, USA, Cat# A-32723 and Cat # A-32740). Nuclear staining was performed with DAPI at 1 µg/ml. Cells were imaged in a Nikon TE widefield fluorescence microscope or a Leica SP8 confocal microscope at the Cell Imaging Core at the University of Utah. Post imaging processing was performed with Imaris Microscopy Image Analysis Software (Oxford instruments).

## Data Availability

All data produced in the present study are available upon reasonable request to the authors

## Acknowledgments

The authors want to express our gratitude to individuals with SCA4, their relatives and care givers. This work was supported by NIH grant R35127253 to SMP. SO received funding from the German Research Foundation DFG (DFG OS 647/1-1). We also acknowledge the Cell Imaging Core at the University of Utah for their use of and assistance with their Nikon and Leica SP8 microscopes. NGS sequencing methods were performed with the support of the DFG-funded NGS Competence Center Tübingen (INST 37/1049-1). JP was supported by the Clinician Scientist program “PRECISE.net” funded by the Else Kröner-Fresenius-Stiftung.

## Supplemental Figures

**Supplemental Fig. 1:**
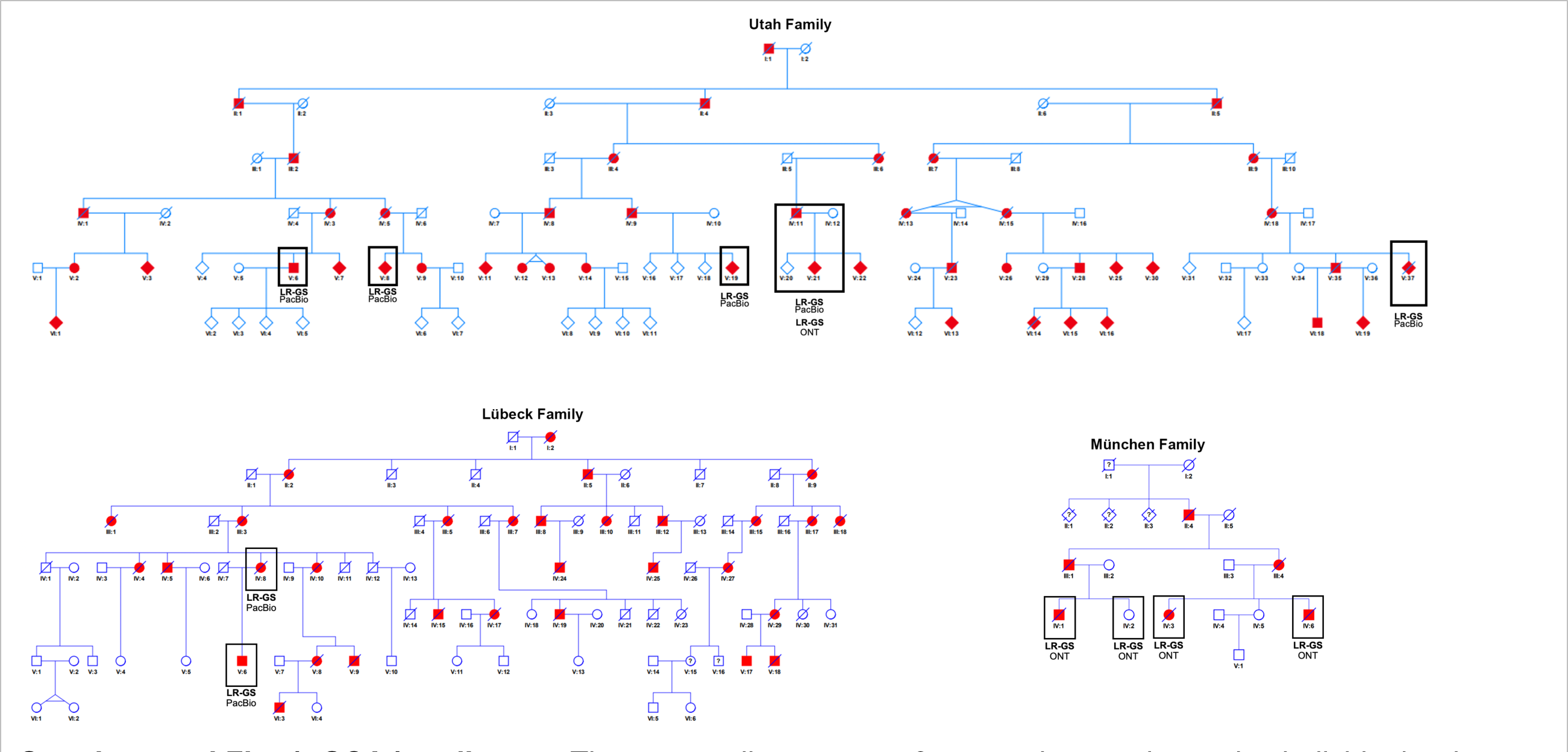
SCA4 pedigrees. The core pedigree spans 6 generations and contains individuals who directly relate back to Utah Mormon Pioneers in the mid 1800’s. The pedigree was constructed based on available genealogical and genetic information (n=526). Red filled symbols denote affected individuals, outlined symbols indicate biological sample collected. The Utah pedigree is altered in its structure including number and order of individuals to maintain anonymity. It represents a subset of the entire pedigree as only 3 of several second generation offspring are depicted. Individuals of a 7th generation have been omitted to further maintain anonymity. Fibroblasts used in the study include V-6, V-8, V-36, and VI-15. Lubeck and Munchen restructured pedigrees are also shown.

**Supplemental Fig. 2:**
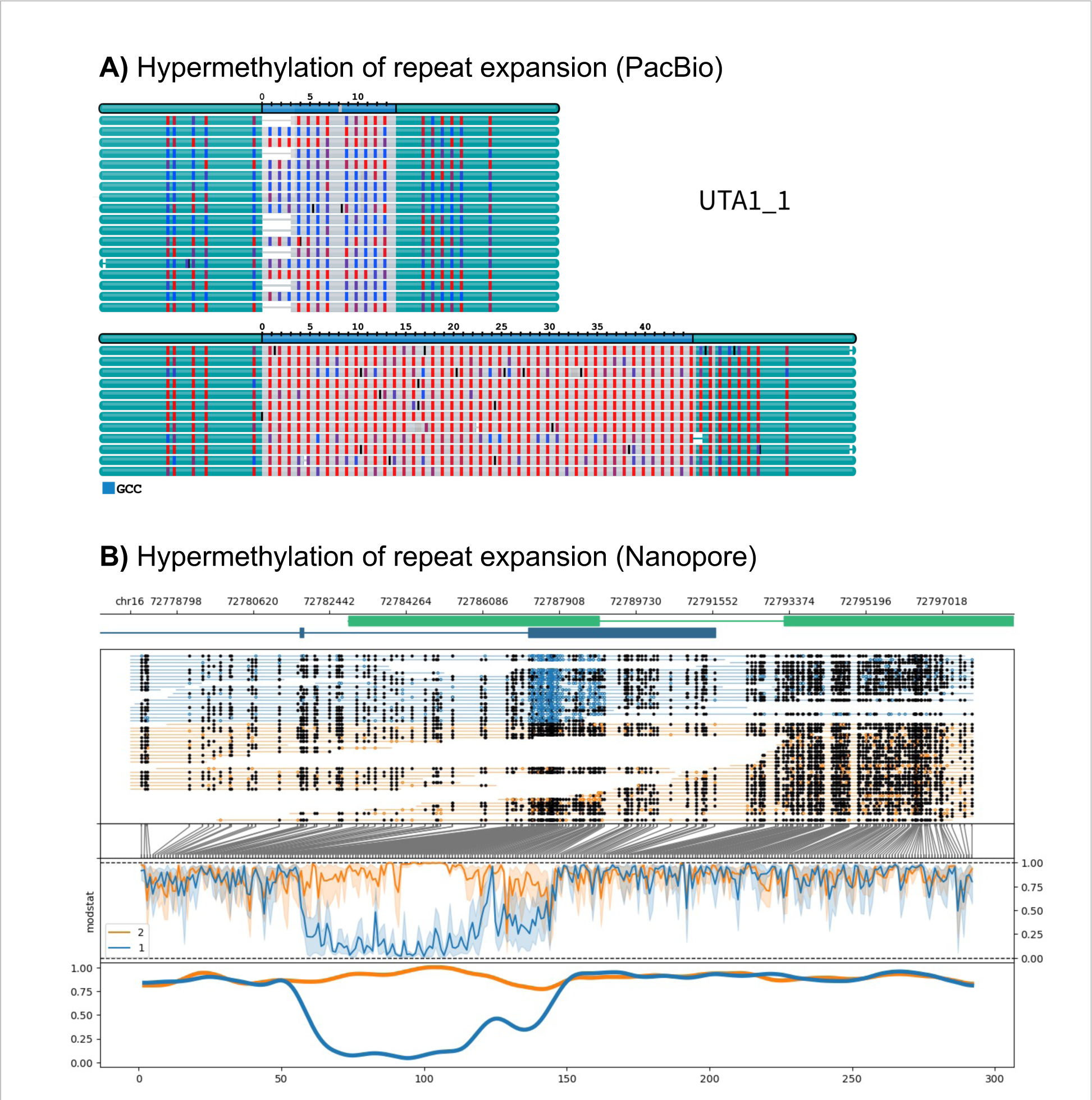
Hypermethylation of the expanded repeat allele. (A) Visualization of methylated cytosines in the repeat expansion by the trgt tool. The expanded allele shows close to 100% methylation in the RE. (B) Visualization of the surrounding region of the RE shows that hypermethylation of the expanded allele (orange) stretches beyond the RE.

**Supplemental Table 1:** Repeat-Expansion discovery. Long-read data from PacBio-SMRT and ONT-Nanopore Sequencing combined with 4 different computational methods was used to identify the RE in *ZFHX3*. Alle Methods reliably identified the RE in affected family members only.

| Seq-Data | Method | Subject Affected | UTA1_1 | UTA1_2 | UTA1_3 | UTA1_4 | UTA2_1 | UTA2_2 | UTA3_1 | UTA1_5 |
| --- | --- | --- | --- | --- | --- | --- | --- | --- | --- | --- |
|  |  |  | + | - | - | + | + | + | + | + |
| PacBio LR | SV calling in de novo assemblies | INS 102bp | + | - | - | + | + | + | + | + |
| PacBio LR | trgt tool using minimap2 alignments |  | + | - | - | + | + | + | + | + |
| ONT LR | SV calling in de novo assemblies | INS 102bp | + | - | - | + | + | + | + | + |
| ONT LR | SV calling with sniffles using minimap2 alignments | INS 99bp | + | - | - | + | + | + | + | + |

**Supplemental Table 2:**
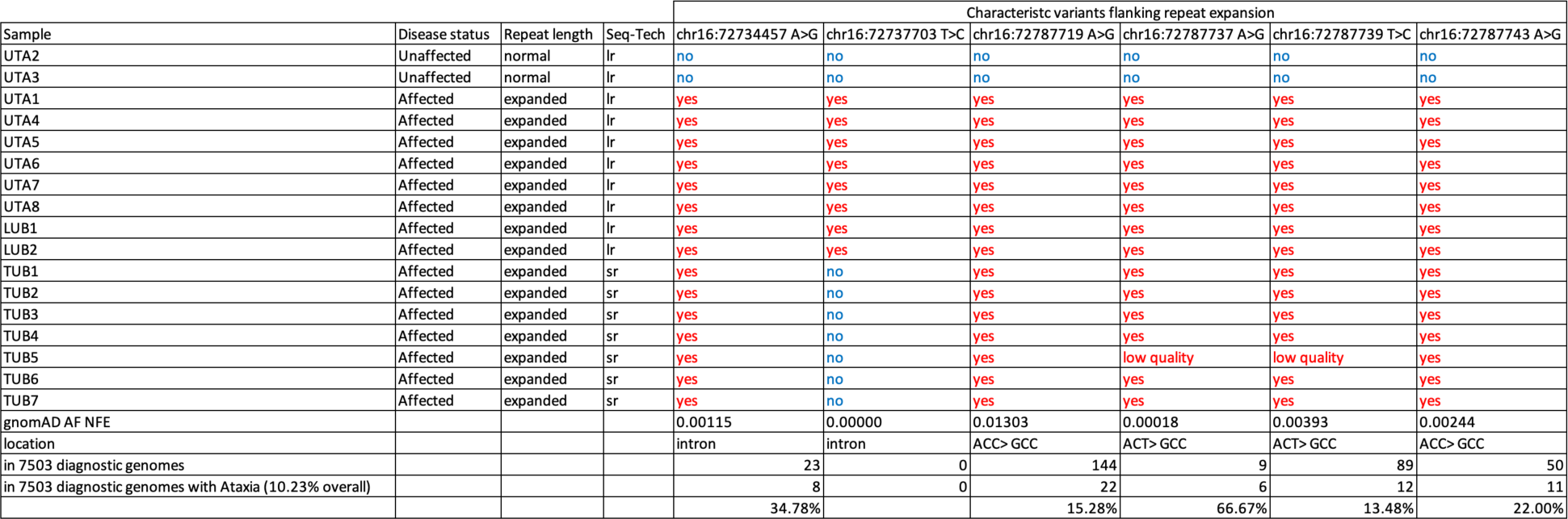
RE-Haplotype characteristics. Six characteristic ultra-rare SNVs reliably identify the RE haplotype in cases from Utah, Lübeck and Tübingen. The 7 samples from Tübingen all are missing the second SNV with a gnomad population frequency of 0. We hypothesize that all samples stem from a single founder and that the Lübeck/Utah haplotype has an additional *de novo* mutation.

